# Comparative accuracy of ChatGPT-o1, DeepSeek R1, and Gemini 2.0 in answering general primary care questions

**DOI:** 10.1101/2025.04.15.25325518

**Authors:** Guerino Recinella, Chiara Altini, Marco Cupardo, Iacopo Cricelli, Lorenzo Maestri

**Affiliations:** Department of Primary Care, AUSL Bologna, Italy; Genomedics SRL, Florence, Italy; Department of Primary Care and Community Medicine AUSL Romagna, Italy

## Abstract

**Objectives:** To evaluate and compare the accuracy and reliability of large language models (LLMs) ChatGPT-o1, DeepSeek R1, and Gemini 2.0 in answering general primary care medical questions, assessing their reasoning approaches and potential applications in medical education and clinical decision-making.

**Design:** A cross-sectional study using an automated evaluation process where three large language models (LLMs) answered a standardized set of multiple-choice medical questions.

**Setting:** From February 1, 2025 to February 15, 2025, the models were subjected to the test questions. For each model, each question was formulated in a new chat session.

Questions were presented in Italian, with no additional instructions. Responses were compared to official test solutions.

**Participants:** Three LLMs were evaluated: ChatGPT-o1 (OpenAI), DeepSeek R1 (DeepSeek), and Gemini 2.0 flash thinking experimental model (Google). No human subjects or patient data were used.

**Intervention:** Each model received the same 100 multiple-choice questions and provided a single response per question without follow-up interactions. Scoring was based on correct answers (+1) and incorrect answers (0).

**Main Outcome Measures:** Accuracy was measured as the percentage of correct responses. Inter-model agreement was assessed through Cohen’s Kappa, and statistical significance was evaluated using McNemar’s test.

**Results:** ChatGPT-o1 achieved the highest accuracy (98%), followed by Gemini 2.0 (96%) and DeepSeek R1 (95%). Statistical analysis found no significant differences (p > 0.05) between the three models. Cohen’s Kappa indicated low agreement (ChatGPT-o1 vs. DeepSeek R1 = 0.2647; ChatGPT-o1 vs. Gemini 2.0 = 0.315), suggesting variations in reasoning.

**Conclusion:** LLMs exhibited high accuracy in answering primary care medical questions, highlighting their potential for medical education and clinical decision support in primary care. However, inconsistencies between models suggest that a multi-model or AI-assisted approach is preferable to relying on a single AI system. Future research should explore performance in real clinical cases and different medical specialties.

## Introduction

Large language models (LLMs) have shown remarkable promise in transforming medical practice. These models have demonstrated the ability to improve diagnostic accuracy, predict disease progression, and support clinical decision-making. ^1 2 3^ Moreover, they have exhibited promising results in various domains, including medical question answering, positioning them to have a significant impact on the healthcare education. ^4^ However, to the best of our knowledge, there are no studies that comprehensively evaluate and compare the reliability and accuracy of LLM-based medical question-answering systems using the “reasoning” technique, such as ChatGPT o1, DeepSeek R1, and Gemini 2.0 flash thinking experimental, in solving general medicine and primary care questions and clinical problems.

Therefore, this study aims to fill this gap by evaluating and comparing the performance of three prominent multi-reasoning system LLMs: ChatGPT o1, DeepSeek R1, and Gemini 2.0 flash thinking experimental, in answering general medical questions.

## Materials and Methods

The study was conducted between February 1 and February 15, 2025. We compared the performance of the ChatGPT o1, DeepSeek-R1 and Gemini 2.0 flash thinking experimental models on a general practice questions. To achieve this, we collected the dataset of general medical questions drawn from the access test for the Italian general medical training course of the year 2023.

We have included all the questions in the official test. These are 100 multiple-choice questions of general medicine and primary care topics. Each question presented five answer options, with only one correct answer. The models analyzed in the study were:

- ChatGPT o1: an advanced language model developed by OpenAI that uses a novel technique called “reasoning” that involves a step-by-step approach to analyze the context, break down the problem, and provide a well-reasoned answer. This approach aims to enhance the model’s ability to engage in structured and logical reasoning, enabling it to tackle complex questions and problems more effectively. ^5^
- DeepSeek R1: an advanced multi-modal language model developed by DeepSeek. The “R1” in the model name denotes its “Reasoning” module, which employs a novel chain-of-thought approach to break down problems, analyze context, and provide well-structured and logical responses. ^6^
- Gemini 2.0 flash thinking experimental model: an advanced experimental large language model developed by Google. This model is designed to leverage a novel “flash thinking” approach, which combines rapid contextual analysis, multi-modal reasoning, and iterative refinement to provide well-structured and logically sound responses to complex questions. ^7^

All models were evaluated using the same set of 100 questions, each presented as an individual prompt within a new chat session to prevent contextual contamination. Each prompt was introduced by the standardized instruction: “For the following question, indicate the correct answer.” Given the multilingual capabilities of all models, the interaction was conducted entirely in Italian, and all questions were formulated accordingly. The complete set of questions is available in the supplemental files.

The chatbot responses were compared with the official test solution. For each correct question, a score of +1 was awarded, and for each incorrect question, a score of zero was awarded. This scoring system was used to fully reflect the official scoring method used to correct the test. Ethics committee approval was not required for this study because no patient data was used.

### Statistical analysis

Statistical analysis was performed with the Statistical Package for the Social Sciences (SPSS) version 25.0. Models performance was evaluated using the following metrics:

- Accuracy: The proportion of correct answers for each LLM
- Cohen’s Kappa: Agreement between the models, accounting for chance agreement.
- McNemar’s Test: To determine whether there was a statistically significant difference in the performance

A p-value of less than 0.05 was considered statistically significant.

## Results

The results of the study showed that the ChatGPT o1 model achieved the highest accuracy, correctly answering 98 out of the 100 general medical questions. The Gemini 2.0 flash thinking experimental model came in second, correctly answering 96 questions. The DeepSeek R1 model had the lowest accuracy, correctly answering 95 questions (Table 1).

**Table 1:** Difference in accuracy between different LLM models.

| Model | Accuracy (%) |
| --- | --- |
| ChatGPT o1 | 98 |
| DeepSeek R1 | 95 |
| Gemini 2.0 flash thinking experimental models | 96 |

The inter-rater reliability, as measured by Cohen’s Kappa, varied considerably across the model pairings. The Kappa value between ChatGPT o1 and DeepSeek R1 was 0.2647, indicating “fair” agreement. Similarly, a “fair” level of agreement was observed between ChatGPT o1 and Gemini 2.0 flash thinking experimental model, with a Kappa value of 0.315. Conversely, the agreement between DeepSeek R1 and Gemini 2.0 flash thinking experimental model was substantially higher, with a Kappa value of 0.651, corresponding to “substantial” agreement.

McNemar’s test did not detect statistically significant differences in any of the comparisons (Table 2).

**Table 2:**
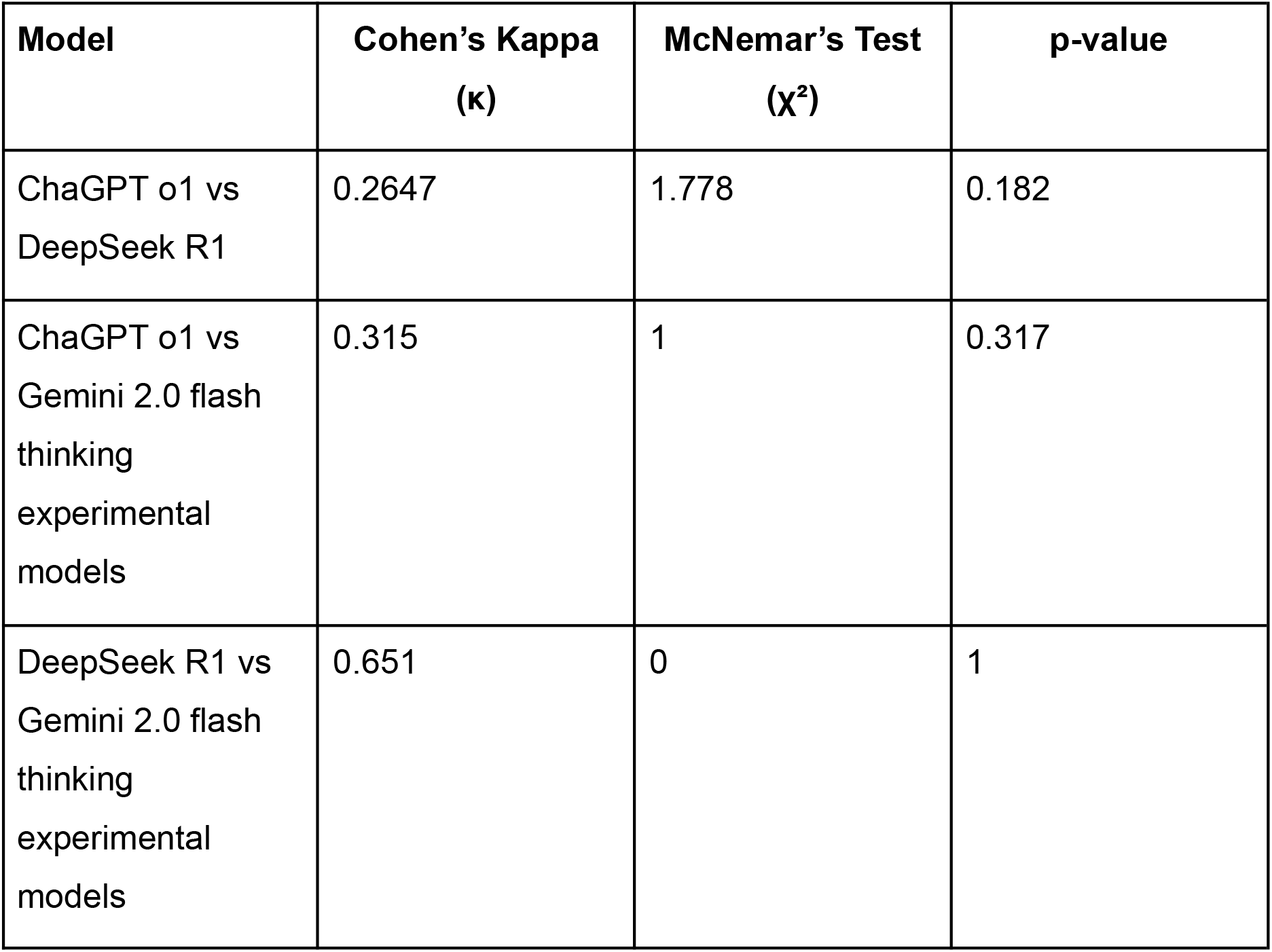
The inter-rater reliability and Comparison Between Models.

| Model | Cohen's Kappa<br>($\kappa$) | McNemar's Test<br>($\chi^2$) | p-value |
| --- | --- | --- | --- |
| ChaGPT o1 vs<br>DeepSeek R1 | 0.2647 | 1.778 | 0.182 |
| ChaGPT o1 vs<br>Gemini 2.0 flash<br>thinking<br>experimental<br>models | 0.315 | 1 | 0.317 |
| DeepSeek R1 vs<br>Gemini 2.0 flash<br>thinking<br>experimental<br>models | 0.651 | 0 | 1 |

Evaluating the errors of individual models, we observe that ChatGPT made two mistakes in the same field: pharmacology. DeepSeek, on the other hand, made errors in questions related to Infectious Diseases (2 errors), Cardiology (2 errors), and Pharmacology (1 error). Gemini, in contrast, made errors in answering questions in Cardiology (2 errors), Infectious Diseases (1 error), and Pharmacology (1 error).

## Discussion

The findings of our study suggest that all three LLM models (ChatGPT o1, DeepSeek R1 and Gemini 2.0 flash thinking experimental) demonstrated a high level of accuracy in answering general medical questions. ChatGPT o1 exhibit the best performance although the differences with Deep Seek R1 and Gemini 2.0 flash thinking experimental model were not statistically significant according to McNemar’s test. This aligns with previous studies that have shown the potential of advanced LLMs, particularly those employing novel reasoning techniques, to support medical education and clinical decision-making. ^8 9 10^

A crucial aspect that emerged from the analysis is the relatively low agreement between the models, as measured by Cohen’s Kappa coefficient. In fact, despite the high individual accuracy, the “fair” agreement between ChatGPT o1 and the other two models and the analysis of individual model errors highlights the potential strengths and weaknesses of each approach. While ChatGPT excelled overall, it exhibited some limitations in the pharmacology domain. In contrast, DeepSeek R1 and Gemini 2.0 both struggled with questions in certain medical domains, such as infectious diseases, cardiology, and pharmacology.

These results suggest that the different approaches to reasoning used by the models, particularly between ChatGPT o1 and the other models, may lead to distinct strengths and weaknesses in addressing specific medical topics. On the other hand, the “substantial” agreement between Deep Seek R1 and Gemini 2.0 suggests that these two models may share more similar features and problem-solving strategies.

However, the results of this study have important implications for the potential use of LLMs in the medical field. While high accuracy is encouraging, the limited agreement between models highlights the need for caution. A single model, even if highly accurate, may not be sufficient to ensure reliable clinical decisions. A multi-model approach or a system where LLMs serve as decision support tools for physicians (rather than substitutes) may be more appropriate.

Our study also had some limitations. It focused on a single type of assessment and a single domain. Moreover, most of the questions on which the models were tested were related to theoretical notions, while questions related to real clinical cases were a small minority. As a result, such a test is hardly representative of everyday clinical reality. It would be useful in future studies to extend the analysis to other types of clinical questions (e.g., clinical cases, open-ended questions) and other medical specialties. It would also be useful to further investigate the reasons behind the limited agreement between models, using a larger and more diverse sample of questions to determine whether it is due to differences in algorithms, training data, or other factors.

In conclusion, our study demonstrates the potential of LLMs in solving general medicine questions but also highlights the need for further research to ensure their safe and effective use in clinical settings.

## Data Availability

All data produced in the present study are available upon reasonable request to the authors.

