## supplemental files for "Comparative accuracy of ChatGPT-o1, DeepSeek R1, and Gemini 2.0 in answering general primary care questions"

**TEST FOR ADMISSION TO THE COURSE OF SPECIFIC TRAINING IN GENERAL MEDICINE IN ITALY. YEAR 2023**

| QUESTION NUMBER | CORRECT ANSWER | QUESTION NUMBER | CORRECT ANSWER | QUESTION NUMBER | CORRECT ANSWER |
|---|---|---|---|---|---|
| 1 | A | 34 | A | 67 | C |
| 2 | E | 35 | E | 68 | C |
| 3 | B | 36 | B | 69 | A |
| 4 | A | 37 | A | 70 | C |
| 5 | E | 38 | A | 71 | B |
| 6 | D | 39 | E | 72 | A |
| 7 | E | 40 | D | 73 | C |
| 8 | A | 41 | A | 74 | B |
| 9 | B | 42 | C | 75 | B |
| 10 | D | 43 | A | 76 | A |
| 11 | C | 44 | B | 77 | C |
| 12 | D | 45 | C | 78 | A |
| 13 | B | 46 | C | 79 | D |
| 14 | B | 47 | D | 80 | B |
| 15 | C | 48 | C | 81 | B |
| 16 | B | 49 | C | 82 | C |
| 17 | D | 50 | D | 83 | A |
| 18 | A | 51 | A | 84 | B |
| 19 | C | 52 | D | 85 | A |
| 20 | C | 53 | C | 86 | A |
| 21 | A | 54 | B | 87 | D |
| 22 | E | 55 | D | 88 | A |
| 23 | B | 56 | A | 89 | D |
| 24 | E | 57 | D | 90 | A |
| 25 | B | 58 | C | 91 | C |
| 26 | A | 59 | D | 92 | D |
| 27 | A | 60 | A | 93 | B |
| 28 | B | 61 | A | 94 | C |
| 29 | B | 62 | B | 95 | B |
| 30 | C | 63 | D | 96 | C |
| 31 | D | 64 | A | 97 | A |
| 32 | B | 65 | B | 98 | B |
| 33 | D | 66 | A | 99 | C |
| | | | | 100 | A |

Each question provides only one correct answer.

Question 1

What is the metabolic cause of Porphyria hepatica acata?

a. Alteration of heme metabolism

b. Alteration of the quaternary structure of Hb

c. Accumulation of amyloid substance

d. Alteration of the Krebs cycle

e. Alteration of the pentose phosphate cycle

Question 2

Which of the following diseases does NOT typically cause interstitiopathy lung?

a. SARS-CoV-2 pneumonia

b. Idiopathic pulmonary fibrosis

c. Lymphangioleiomyomatosis

d. Hypersensitivity pneumonitis

e. Streptococcus pneumoniae pneumonia.

Question 3

Which of the following criteria allows a diagnosis of diabetes mellitus to be made?

a. Fasting blood glucose from venous sampling greater than or equal to 110 mg/di on two occasions

b. Fasting blood glucose from venous sampling greater than or equal to 126 mg/dl on two occasions

c. Occasional detection of blood glucose greater than 150 mg/dl

d. Occasional detection of blood glucose greater than 180 mg/dl

e. HbAlc value of less than 48 mmol/mol

Question 4

What is community pneumonia?

a. A pneumonia that is contracted outside of hospitals and residential facilities

b A pneumonia that is contracted only within hospital communities

c. A pneumonia with viral etiology

d. A pneumonia of unknown etiology

e. A pneumonia affecting an entire community

Question 5

Virchow-Troisier's lymph node occurs:

a. at the right inguinal level

b. at the right laterocervical level

c. in the left popliteal fossa

d. in the mediastinum

e. at the left supraclavicular level

Question 6

Which of the following antihypertensive drugs is indicated in pregnancy?

a. Enalapril

b. Hydralazine

c. Doxazosin

d. Alpha-methyldopa

e. Valsartan

Question 7

All of the following examinations are included in the population screening campaign

for early detection of cancer in the Italian population, except:

a. mammography for breast cancer in women

b. fecal occult blood detection for colorectal carcinoma in men

c. fecal occult blood test for colorectal carcinoma in women

d. pap smear test for cervical carcinoma in woman

e. PSA for carcinoma of the prostate in men

Question 8

A 55-year-old woman has been complaining of dysphagia mainly from liquid foods for 2 months. In suspicion of achalasia, which of the following is the test that allows diagnosis?

a. Esophageal manometry

b. Chest x-ray

c. CT chest and abdomen with contrast medium

d. EGDS

e. Abdominal ultrasonography

Question 9

The most common causes of dementia are:

a. deficiency of thiamine, niacin, vitamin B12

b. Alzheimer's disease, vascular dementia, Lewy body dementia

c. intoxication by lead, mercury or other heavy metals

d. neurositilide, toxins

frontotemporal dementia (Pick's disease), normotensive e. hydrocephalus dementia

Question 10

Influenza vaccination is:

a. recommended exclusively for women in the 1st trimester of pregnancy

b. recommended exclusively to women in the II trimester of pregnancy

c. recommended exclusively to women in the 3rd trimester of pregnancy

d. recommended to women who are pregnant at the beginning of the epidemic season

e. contraindicated in pregnancy

Question 11

Chvostek's sign is indicative of:

a. meningeal irritation

b. hyperkalemia

c. hypocalcemia

d. possible cholecystitis

e. inflammation of the ileo-psoas muscle

Question 12

Which of these femoral or vertebral T-score values is indicative of

osteoporosis?

a. between -2 and -1

b. between -1 and + 2.5

c. > + 2,5

d. <- 2,5

e . between -1 and 0

Question 13

Prescription for treatment of gynecological Chlamydia trachomatis infections should be given:

a. only to the person presenting the infection

b. to the person presenting the infection and the sexual partner

c. with at least two courses of antibiotic therapy

d. with at least two antibiotics in combination

e. with probiotics

Question 14

Which of the following is NOT an antiepileptic drug?

a. Sodium valproate

b. Rasagiline

c. Lacosamide

d. Levetiracetam

e. Diazepam

Question 15

The use of which of the following classes of drugs may result in an increased risk of developing Clostridium difficile infection?

a. Loop diuretics

b. Potassium-sparing diuretics

c. Proton pump inhibitors

d. MAO inhibitors

e. Antihistamines

Question 16

Which direct oral anticoagulant is contraindicated for creatinine clearance < 30 ml/min in nonvalvular atrial fibrillation?

a. Apixaban

b. Dabigatran

c. Edoxaban

d. Rivaroxaban

e. Warfarin

Question 17

Which rare disease should be suspected in patients presenting with chronic hemolytic anemia, atypical arterial and/or venous site thrombosis, and marked asthenia?

a. Macrophage activation syndrome

b. Autoinflammatory syndrome

c. Alpha-thalassemia

d. Nocturnal paroxysmal hemoglobinuria (PNH)

e. Transthyretin amyloidosis (ATTRv)

Question 18

Which people are most at risk for pneumococcal diseases such as pneumonia and meningitis?

a. Children and the elderly, particularly if burdened by concomitant diseases

b. Ecological workers

c. The employees of public administrations

d. School personnel

e. Canteen staff

Question 19

In secondary bacterial pneumonia which germ represents by far the most frequent etiologic agenta. Pseudomonas aeruginosa

b. Enterobacter cloacae

c. Streptococcus pneumoniae

d. Haemophilus influenzae

e. Respiratory syncytial virus

Question 20

COPD:

a. is defined as a completely reversible airflow limitation

b. recognizes cigarette smoking as the sole risk factor

c. and . is stratified according to GOLD criteria

d. is determined by a chronic increase in FEVI

is a disease with declining incidence over the past 20 years

Question 21

Telemonitoring in the heart failure patient:

a. enables patients to remotely provide information about their health status so to support and optimize their care

b. lacks evidence of efficacy and safety

c. induces reduced therapeutic adherence

d. can only be implemented in patients with PM/ICD

e. induces excessive emotional stress

Question 22

Which of the following is the definition of paradoxical dysphagia?

a. Difficulty in sensing tastes

b. Difficulty formulating words

c. Difficulty looking to the left

d. Inability to smell smells

e. Difficulty swallowing liquids

Question 23

The ketogenic diet is strongly hypocaloric and is contraindicated in all of the following conditions except:

a. liver failure

b. obesity and dyslipidemia

c. chronic renal failure

d. heart failure

e. diabetes mellitus type 1

Question 24

Which of these statements is true?

a. Fondaparinux belongs to the class of direct oral anticoagulant drugs and can be administered every 24 hours

b. The desired INR value for patients with venous thromboembolism is between 3 and 4

c. Rivaroxaban is a direct thrombin inhibitor and does not require monitoring with laboratory tests

d. Daily use of vascular compression stockings in the acute phase of venous thromboembolism increases the risk of developing postphlebitic syndrome

e. Direct oral anticoagulants are administered at a fixed dosage and do not require laboratory monitoring of anticoagulant activity

Question 25

What is the mechanism of action of sacubitril-valsartan?

a. Antagonism of aldosterone and inhibition of the sodium-glucose pump

b. Angiotensin antagonism and inhibition of neprilysin

c. Antagonism of noradrenaline and inhibition of the converting enzyme of

angiotensin

d. Angiotensin antagonism and phosphodiesterase inhibition

e. AT1 receptor antagonism and inhibition of the renin-angiotensin system

Question 26

In the early stage of diabetic nephropathy we can find:

a. glomerular hypertrophy and increased filtration rate

b. decreased glomerular filtrate values with eGFR $< 60$ ml/min/1.73 sq m

c. always changes in urinary sediment

d. proteinuria with values $> 3$ g/24 h

e. positive leukocyte esterase

Question 27

Which of these signs is NOT present in Bernard-Horner syndrome?

a. Lateral deviation of gaze

b. miosis

c. Anhidrosis

d. Eyelid ptosis

e. Endophthalmos

Question 28

The presence of chronic renal failure changes the risk profile cardiovascular:

a. only in patients with other risk factors

b. increases it even in the absence of other risk factors

c. is never a risk factor

d. only for filtrate values below 30 ml/min/1.73 sq m

e. only for filtrate values less than 15 ml/min/1.73 sq m

Question 29

In a patient with nonvalvular atrial fibrillation with CHA2DSz-VASc 3 and HASBLED 4, in the absence of other contraindications, the best strategy for prophylaxis of systemic thromboembolism is represented by:

a. no anticoagulant treatment because the hemorrhagic risk outweighs the ischemic

b. anticoagulant treatment with direct-acting oral anticoagulants and frequent follow-up to intervene on modifiable bleeding risk factors

c. anticoagulant treatment with dicumarolics

d. anticoagulant treatment with low molecular weight heparin

e. treatment with oral anticoagulants only after correction of risk factors of modifiable bleeding

Question 30

In heart failure, SGLT2-inhibitor drugs:

a. were shown to be effective if the patient also had type 2 diabetes

b. reduced hospitalization and mortality from heart failure only in

patients who also had type 2 diabetes

c. reduce hospitalization and mortality from heart failure

regardless of the presence of diabetes

d. reduce lo oraliarica i the mortality from heart failure only so

e. are second-choice drugs in the therapy of heart failure

Question 31

The main treatment in Pickwick syndrome is represented by:

a. benzodiazepines

b. methylphenidate

c. beta-blockers

d. noninvasive mechanical ventilation

e, amitriptyline

Question 32

Administration of the sacubitril-valsartan combination in patients with heart failure results in:

a. significant increase in the incidence of cardiovascular death

bo significant reduction in cardiovascular death and hospitalization for heart failure

c. significant reduction in cardiovascular death but not hospitalization for heart failure

d. significant reduction of hospitalization for heart failure, but not of cardiovascular death

e. significant reduction in cardiovascular death, but only in association with enalapril-hydrochlorothiazide

Question 33

In a patient who developed a fever one week after a tick bite, the most appropriate test is:

a. Widal-Wright serodiagnosis

b. High-sensitivity PCR

c. Rheumatoid factor

d. Weil-Felix serodiagnosis

e. Parasitological stool examination

Question 34

In which of the following conditions is reduced thyroid uptake NOT present on scintigraphy?

a. Graves' disease

b. Subacute thyroiditis

c. Silent thyroiditis

d. Amiodarone thyroiditis

e. Factitious thyrotoxicosis

Question 35

An EchocolorDoppler examination in a 59-year-old subject shows a 20% asymptomatic carotid stenosis. What therapeutic indication is correct to this patient?

a. Vascular surgery for carotid unblocking

b. Oral anticoagulant therapy

c. Study of morphology and extent of stenosis with an AngioTC examination

d. Referral to interventional radiologist for carotid artery stenting surgery

e. Correction of risk factors with appropriate lifestyle and medical therapy

Question 36

The pathogen called Helicobacter pylori is:

a. Gram-positive cocci

b. Gram-negative bacillus

c. protozoan

d. virus

e. mycete

Question 37

A positive Lasegue maneuver indicates:

a. L5-$1 nerve root irritation

b. severe coxarthrosis

c. sub-acromial conflict syndrome

d. suspected meniscal injury

e. possible anterior cruciate ligament injury

Question 38

Which of these features is NOT found in patients with nephrolithiasis?

a. Hypocalciuria

b. Hyperuricosuria

c. Primary hyperparathyroidism

d. Hyperoxaluria

e. Hypocitraturia

Question 39

COVID-19 is a disease:

a. of the upper airway

b. of the lower airway

c. of the kidney

d. of the limbs

e. multi-organ

Question 40

What is meant by high normal blood pressure?

a. Systolic pressure between 160 and 169 mmHg or diastolic pressure between 100 and 109 mmHg

b. Systolic pressure between 140 and 159 mmHg or diastolic pressure between 90 and 99 mmHg

c. Systolic pressure between 120 and 129 mmHg or diastolic pressure between 80 and 84 mmHg

d. Systolic pressure between 130 and 139 mmHg or diastolic pressure between 85 and 89 mmHg

e. Systolic pressure >180 mmHg or diastolic pressure > 110 mmHg

Question 41

The mechanism of action of warfarin is as follows:

a. vitamin K antagonist

b. vitamin D3 antagonist on osteoblasts

c. vitamin B12-intrinsic factor complex antagonist

d. vitamin B8 (biotin) antagonist

e. folic acid antagonist

Question 42

In a patient who has acutely developed symptoms characterized by right flank pain radiating to the groin, associated with nausea and vomiting, the first-level diagnostic test to be requested is:

a. CT abdomen with and without contrast medium

b. uroflowmetry

c. abdominal ultrasound

d. renal scintigraphy

e. ECG in emergency

Question 43

Ankle Brachial Index (ABI) is considered definitely pathological for a value:

a. < 0.9

b. 1.1

c. 1

d. 1.2

e. 1.25

Question 44

Which of these classes of drugs most frequently causes constipation?

a. Penicillins

b. Opioid analgesics

c. Benzodiazepines

d. Statins

e. Biguanides

Question 45

The diagnosis of COPD is made by:

a. six minute walking test

b. lung ultrasound

c. spirometry

d. lung plethysmography

e. polysomnogram

Question 46

In patients with chronic heart failure with reduced ejection fraction, which of these drug classes is NOT recommended because of an increased risk of adverse effects (lower extremity edema, dyspnea, edema

a. Beta-blockers

b. Glyflozines

c. Calcium channel blockers

d. ACE inhibitors

e. ARNI (sacubitril-valsartan)

Question 47

Which class of antibiotics can promote the development of carbapenemase-producing Klebsiella strains?

a. Macrolides

b. Betalactams

c. Carbapenemases

d. Quinolonics

e. Glycopeptides

Question 48

Which among the following clinical conditions can be attributed to side effect of amiodarone?

a. Secondary hyperaldosteronism

b. Inappropriate ADH augmentation syndrome

c. Hyper- or hypothyroidism

d. Iatrogenic diabetes

e. Hyperprolactinemia syndrome

Question 49

In Graves' disease, which of the following makes one opt in favor of radiometabolic therapy?

a. Pregnancy

b. Active ophthalmopathy

c. Comorbidities that increase surgical risk

d. Suspected malignancy

e. Presence of high-volume nodules

Question 50

Environmental pollutants are responsible for organ damage for all of the following diseases, except:

a. ischemic heart disease

b. COPD

c. lung cancer

d. celiac disease

e. lower airway infections

Question 51

The mechanism of renal action of the drugs defined as SGLT2-inhibitors is achieved:

a. by inhibiting the sodium-glucose co-transporter in the proximal convoluted tubule

b. by inhibiting the effect of angiotensin in the proximal convoluted tubule

c. by inhibiting the effect of angiotensin in the distal tubule

d. by inhibiting the receptor for ACE (Angiotensin Converting Enzyme)

e. by inhibiting the receptor for vitamin D3

Question 52

Which statement about ventilator-associated pneumonia (VAP) is correct?

a. The endotracheal tube and the need for suction are not risk factors

b. Ventilator-associated pneumonia infection correlates with a shorter period of hospital and ICU stay

c. Empirical antibiotic treatment does not vary by the presence of risk factors for multi-resistant pathogens

d. Multi-resistant pathogens are associated with higher mortality than non-multi-resistant pathogens

e. Clinical manifestations do not include increased frequency respiratory

Question 53

In immunocompromised and immunosuppressed individuals, influenza and pneumococcal vaccines:

a. cannot be co-administered

b. are contraindicated

c. can and should be administered

d. can only be administered in the female sex

e. can only be administered in children

Question 54

Which of the following procedures is NOT recommended in the evaluation of thyroid nodule?

a. Ultrasound

b. Thyroglobulin assay

c. TSH assay

d. Thyroid scintigraphy if TSH is reduced

e. Accurate history and objective examination

Question 55

Which of the following is NOT an indication for performing esophagogastroduodenoscopy?

a. Upper gastrointestinal bleeding

b. Removal of esophageal foreign body

c. Placement of gastrostomy

d. Ileal stenosis

e. Palliative treatment of duodenal stenosing neoplasm

Question 56

Which among these laboratory changes should be considered as a "red flag" for Paroxysmal Nocturnal Hemoglobinuria (PNH)?

a. LDH > 800 mU/ml

b. AST > 5 times the maximum normal value

c. Cholesterolemia > 250 mg/ml

d. PCHE (Pseudo Cholinesterase) < 2000 U/L

e. Ferritinemia < 15 ng/ml

Question 57

Which of these hypoglycemic drugs has been shown to improve prognosis in patients with chronic heart failure with reduced ejection fraction?

a. Metformin

b. Sitagliptin

c. Insulin degludec

d. Empagliflozin

e. Pioglitazone

Question 58

Which of the following pathological conditions of the respiratory system is NOT typically associated with obstructive atelectasis?

a. Small cell lung carcinoma

b. Foreign body inhalation

c. Pleural empyema

d. Mucus plugs

e. Squamous cell lung carcinoma

Question 59

Which of these active ingredients does NOT find indication in the treatment of delirium in a palliative care setting?

a. Haloperidol

b. Chlorpromazine

c. Risperidone

d. Allopurinol

e. Midazolam

Question 60

CRT (Cardiac Resynchronization Therapy):

a. is recommended to reduce morbidity in patients with heart failure at moderately reduced ejection fraction (HFmrEF), including those with atrial fibrillation, who have an indication for pacing ventricular pacing and high-grade AV block regardless of NYHA class and

by QRS duration

b. is proposed only in BBdx

c. is indicated in pulmonary arterial hypertension

d. increases sympathetic autonomic nervous system stimulation

e. may cause supraventricular arrhythmias

Question 61

The recommended treatment for adults and adolescents with latent, primary, secondary, or early syphilis is:

a. benzathine penicillin G intramuscularly

b. amoxicillin per os

c. cefuroxime intramuscularly

d. clarithromycin per os

e. cotrimoxazole per os

Question 62

Which eGFR (estimated glomerular filtrate) value identifies the patient with stage III chronic renal failure?

a. < 15 ml/min/1.73 sq m

b. 30-59 ml/min/1.73 sq m

c. 40-75 ml/min/1.73 sq m

d. 60-89 ml/min/1.73 sq m

e. 15-29 ml/min/1.73 sq m

Question 63

What is the minimum value of bacterial load to consider a urine culture definitely positive?

a. 100 CFU/ml

b. 1,000 CFU/ml

c. 10,000 CFU/ml

d. 100,000 CFU/ml

e. 1,000,000 CFU/ml

Question 64

Flu vaccine should NOT be administered:

a . to infants under six months

b. to adults

c. to pregnant women

d. to the elderly

e. to institutionalized individuals

Question 65

Cheyne-Stokes breathing:

a. c. is characterized by an apnea phase in which pO2 increases and pCO2 decreases

b. is present in patients with advanced heart failure and is associated with low cardiac output is always associated with increased cardiac output

d. is always absent in patients with heart failure

e. is caused by increased sensitivity of the breathing centers to arterial pCO2 levels

Question 66

Nirmatrelvir/ritonavir is a:

a. antiviral

b. antibacterial

c. antiprotozoal

d. disinfectant

e. antifungal

Question 67

Which of the following ultrasound criteria is considered highly specific for suspecting malignancy of a thyroid nodule?

a. Intra-nodular vascularization

b. Macrocalcifications

c. Microcalcifications

d. Hypoechogenicity

e. Spongiform appearance

Question 68

The best treatment of F3 esophageal varices with red spots is:

a. octreotide

b. propranolol

c. esophageal varices ligation and prophylaxis with non-selective beta-blockers

d. esophageal varices ligation and prophylaxis with selective beta-blockers

e. ACE inhibitors and sartans

Question 69

Currently in the genus Legionella are classified:

a. 48 species comprising 70 serogroups

b. 4 species

c. 15 species

d. 20 species

e . 2 species

Question 70

In a naive third-degree hypertensive patient, which drug should be administered during screening to rule out secondary forms of hypertension?

a. ACE inhibitor

b. Sartan

c. Calcium channel blocker

d. Beta-blocker

e. Alpha-methyldopa

Question 71

Which of the following features is typical of bronchial asthma?

a. Restrictive ventilatory deficit

b. Bronchial hyperresponsiveness

c. Impaired CO diffusing capacity

d. Irreversible obstruction

c. Interstitial thickening

Question 72

Which of the following is NOT an antiviral vaccine?

a. Anti-tetanus

b. Anti-morbillus

c. Anti-rubella

d. Anti-parotitis

e. Anti-varicella

Question 73

Bronchial asthma therapy:

a. makes use of antileukotrienes in the early stages of disease, as they are more effective than inhaled steroids

b. is independent of symptom severity and number of flare-ups

c. makes use of long-acting beta2-agonists and inhaled glucocorticoids, also in combination

d. does not include omalizumab in patients with frequent flare-ups

e. involves the use of low doses of systemic corticosteroids in the early stages of disease, to reduce its progression

Question 74

Moderately reduced ejection fraction heart failure (HFmrEF) indicates an ejection fraction:

a. of 30-40%

b. of 41-49%

c. of 40-50%

d. > 50%

e. < 30%

Question 75

Which of these clinical pictures should suggest chronic heart failure ?

a. Fever, dyspnea and polypnea, polyarthralgias

b. Exertional dyspnea, orthopnea, declivous edema, jugular turgor

c. Unilateral pretibial and perimalleolar edema, positive Homans' sign

d. Osler's nodules, Janeway's palm spots, fever

e. Tachypnea, precordial pain and precordial auscultatory finding of rubs

Question 76

Hypertensive arteriolar nephrosclerosis is a progressive renal disease caused by:

a. poorly controlled arterial hypertension of old date

b. occasional hypertensive spikes in hypertensive usually well controlled with ongoing therapy

c. poorly controlled systolic only hypertension of old date

d. poorly controlled diastolic only hypertension of old date

e. well controlled systolic-diastolic only hypertension, but of old date

Question 77

In what clinical condition can Janeway's spots palm or plantar be present?

a. Constrictive pericarditis

b. Discoid lupus

c. Bacterial endocarditis

d. Petechial typhus

e. Pellagra

Question 78

The pathogen of diphtheria is:

a. Gram-positive bacillus

b. Gram-negative bacillus

c. Gram-positive coccus

d. protozoan

e. single-stranded DNA virus

Question 79

Which of the following is NOT a pro-inflammatory cytokine?

a. Interferon

b. Interleukin 2

c. Granulocyte growth factor

d. Ferritin

e. Tumor necrosis factor alpha

Question 80

In a patient with nonvalvular atrial fibrillation being treated with apixaban, the reference formula for estimating renal function is given by:

a. MDRD

b. creatinine clearance calculated by Cockroft-Gault formula

c. BIS-1

d. CKD-EPI

e . BIS-2

Question 81

Which of these diseases is NOT associated with vesiculobullous or pustular rashes?

a. Primary infection with Herpes Simplex Virus (HSV)

b. Scarlet fever

c. Hand-foot-mouth syndrome

d. Varicella

e. Smallpox

Question 82

What does the presence of asterisks indicate in a patient?

a. Parkinson's disease

b. Atherosclerotic Parkinsonism

c. Hepatic encephalopathy

d. Polyneuritis

e. Radicular compression

Question 83

In patients with chronic ischemic heart disease who present with residual dyspnea despite first-line medical therapy, which of these drugs can be used to improve symptoms?

a. Ranolazine

b. Thiazide diuretic

c. Rivaroxaban

d. Furosemide

e. Atorvastatin

Question 84

The diagnosis of COPD on spirometry examination is confirmed by the VEMS/CVF (Maximum Expiratory Volume at First Second/Forced Vital Capacity) ratio:

a. less than 0.95 post-bronchodilation

b. less than 0.70 post-bronchodilation

c. less than 0.75 post-bronchodilation

d. less than 0.80 post-bronchodilation

e. less than 0.85 post-bronchodilation

Question 85

The posterior drawer maneuver is used to suspect possible injuries of:

a. posterior cruciate ligament

b. medial meniscus

c. goosefoot tendon

d. anterior cruciate ligament

e. medial collateral ligament

Question 86

Influenza vaccine offers specific protection:

a. exclusively for influenza epidemic disease

b. for all acute respiratory diseases

c. only for bacterial diseases

d. only for parasitic diseases

e. only for chronic diseases

Question 87

Which of the following systemic diseases is NOT a cause of skin itching?

a. Chronic renal failure in the uremic phase

b. Carcinoid syndrome

c. Primary biliary cholangitis

d. Idiopathic pulmonary fibrosis

e. Scleroderma

Question 88

In what typical way does the flu differ from the common cold?

a. Abrupt onset

b. Gradual onset

e. Abdominal pain

d. Lymphadenomegaly

e. Pancreatitis

Question 89

Which of these active ingredients is NOT indicated in the treatment of hepatitis B:

a. Lamivudine

b. Entecavir

c. Telbivudine

d. Lamotrigine

e. Tenofovir

Question 90

Influenza viruses are RNA viruses of the family Orthomyxoviridae; there are:

a. four main types

b. two main types

c. five main types

d. three main types

e. seven main types

Question 91

Metformin therapy in diabetes mellitus requires care in checking periodically:

a. adrenal function (cortisol)

b. pancreatic function (amylase, lipase)

c. renal function (creatinine clearance)

d. ketonemia and ketonuria

e. lipid metabolism

Question 92

What does the ultrasound finding of no change in the caliber of the vein Inferior Cava Inferior with respiratory excursions indicate?

a. Physiologic finding

b. Aortic stenosis

c. Rheumatic fever

d. Heart failure

e. Aneurysm of the thoracic aorta

Question 93

Statins reduce endogenous cholesterol synthesis by inhibiting competitively the activity of which enzyme?

a. Succinyl-CoA-synthetase

b. HMG-CoA-reductase

c. Alpha-ketoglutarate-dehydrogenase

d. Malate-dehydrogenase

e. Fumarate-hydratase

Question 94

How can COVID-19 vaccines be administered compared with influenza vaccines?

a. At least 4 weeks apart

b. At least 1 month apart

c. Simultaneously or at any time before or after

d. At least 2 weeks apart

e. At a minimum distance of 1 day

Question 95

How does the protruded tongue behave in right 12th cranial nerve palsy?

a. Deviates to the left

b. Deflects to the right

c. It does not deviate

d. The tongue cannot be protruded

e. Deviates downward

Question 96

Which of these drugs can cause a photosensitivity reaction if the patient sỉ exposes to the sun during treatment?

a. Ramipril

b. Allopurinol

c. Doxycycline

d. Cholecalciferol

e. Amoxicillin

Question 97

Ramsay Hunt syndrome (herpes zoster oticus) affects:

a. the geniculate ganglion with impairment of the VIII cranial nerve

b. the trigeminal nerve

c. the femoro-crural nerve

d. the sciatic nerve

e. the ophthalmic nerve

Question 98

Which group of examinations should be required for evaluation of organ damage in hypertension?

a. Echocardiogram, fundus oculi, CT brain without contrast

b. Echocardiogram, fundus oculi, Doppler TSA

c. Echocardiogram,,fundus oculi, Doppler renal arteries, B-mode ultrasound urinary tract

d. Echocardiogram, fundus oculi, Doppler renal arteries, B- mode ultrasound urinary tract, CT encephalon without contrast

a. Echocardiogram, fundus oculi, renal artery Doppler, B-mode ultrasound urinary tract, encephalon CT scan without contrast, myocardial scintigraphy

Question 99

Treatment with beta-blockers in patients with heart failure:

a. reduces symptoms of heart failure, but does not improve clinical status

b. reduces hospitalizations, but not the risk of death

c. reduces the risk of death and the combined risk of death or hospitalization

d. reduces heart rate in a manner that is not proportional to selectivity for beta-adrenergic receptors

e. reduces the consumption of diuretic drugs and vasodilators

Question 100

What is the prevalence of chronic heart failure in the general population in patients older than 70 years?

a. > 10%

b. 1-2%

c. 5%

d. 4%

e. 3%
